# Healthcare professionals’ perspectives on the challenges faced by parents of children with autism and recommendations to address them: A qualitative study

**DOI:** 10.1101/2025.11.13.25340157

**Authors:** Karen KY Ma, Anne-Marie Burn, Oscar WH Wong, Olivia Choi, Sandra SM Chan

**Affiliations:** Department of Psychiatry, Faculty of Medicine, The Chinese University of Hong Kong, Hong Kong SAR; MRC Epidemiology Unit, School of Clinical Medicine, University of Cambridge, United Kingdom; Department of Psychiatry, School of Clinical Medicine, University of Cambridge, United Kingdom

**Keywords:** Parents, Caregivers, Stress, Autism spectrum disorders, Healthcare professionals

## Abstract

**Objective:** To explore healthcare professionals’ (HCPs) perspectives on the challenges and needs of parents caring for children with autism spectrum disorder (ASD).

**Design:** In-depth individual and group interviews were conducted with 16 HCPs with over 10 years’ experience working closely with children with autism and their families. HCPs interviewed provided tertiary specialist clinical services for ASD across a range of roles, including child psychiatrists, psychologists, psychiatric nurses, a social worker, an occupational therapist and a hospital-based teacher. Data was coded inductively and analysed using reflective thematic analysis.

**Setting:** A university-affiliated tertiary hospital in Hong Kong.

**Results:** Three themes were generated: (1) Emotional strain and its negative impact on parenting practices, (2) Misunderstanding about ASD and its management, and (3) Acknowledgement and acceptance of the ASD diagnosis. HCPs observed that challenges faced by parents and caregivers of ASD were often shaped by misinformation and misinterpretation of child behaviours and needs, as well as unrealistic expectations based on stereotypes and norms. HCPs provided recommendations for parents and caregivers, highlighting the need to strengthen relational skills to connect with their children, take their child’s perspective, and show appreciation and affection for their child’s strengths.

**Conclusions:** Findings from this study highlighted the need for psychoeducation and identified the potential loci and timing for intervention to better support parents and caregivers of ASD in overcoming challenges and building stronger relationships with their children. This has important practical implications for mitigating bidirectional mutual perpetuation of parental stress and child behavioural and emotional problems.

*Key messages:* What is already known on this topic

- Parents and caregivers of children with autism face challenges around the diagnosis, including experiencing a range of emotions, coping with children’s behaviour problems, and dealing with stigma.
- Previous studies on caregiving for children with autism have mostly centred on parents’ and caregivers’ perspectives, but they may not always have the skills and experiences to recognise exactly what they are struggling with. What this study adds

- HCPs observed that misunderstandings about ASD and its management is a key challenge faced by parents and caregivers, which was shaped by misinformation, misinterpretation of child needs, and misconstrued behavioural reinforcement.
- Key recommendations from HCPs for parents and caregivers to overcome challenges include improving relational skills, cultivating curiosity in parenting, setting consistent boundaries and limits, and prioritising self-care. How this study might affect research, practice, or policy

- Psychoeducation can be incorporated in clinicians’ routine practice to better support parents and caregivers of ASD and improve family wellbeing and quality of life.
- Future research should explore whether strengthening relational skills can improve parent-child interactions and child behavioural outcomes, and enhance family wellbeing and quality of life.

## Introduction

Autism spectrum disorder (ASD) is a neurodevelopmental disorder characterised by social communication difficulties and restricted, repetitive behaviours that affect social, emotional, and cognitive functioning. ASD persists across the lifespan, with many individuals experiencing functional impairments and behavioural challenges [1]. These difficulties have a profound impact not only on children, but also on their parents and caregivers, who serve as their primary support network and face numerous challenges in raising their children [2]. Accordingly, research consistently reports elevated levels of depression and anxiety among parents of children with ASD [3], which must be addressed to reduce the bidirectional perpetuation of parental stress and child behavioural and emotional difficulties [4].

Parents and caregivers of children with ASD face a range of unique challenges through the pathway of care, from pre-diagnosis, diagnosis and access to care, to post-diagnosis [2,5]. Before receiving a formal ASD diagnosis, parents often struggle to understand their child’s behavioural and communication difficulties, while becoming increasingly aware of atypical development [6]. During the diagnostic process, delays in assessment, inadequate explanations of diagnosis, and parents’ own emotional responses are common challenges amongst parents of children with ASD [7]. After receiving the formal diagnosis, parents navigate healthcare and educational systems [8,9], cope with their children’s challenging behaviours [10], and deal with stigma, judgment, rejection, and exclusion [11,12]. Intra- and interpersonal factors such as coping styles, self-efficacy, marital and coparenting quality, and family dynamics can also affect parental stress [13,14].

While caregiving challenges may differ according to healthcare systems and service availability, parents consistently report emotional strain and children’s behavioural problems as key difficulties across contexts [15,16]. This suggests that while broader structural or cultural factors may play a role, interpersonal and psychosocial factors play a central role in shaping everyday challenges. However, as parents and caregivers may not always recognise how their own psychological processes shape their responses and coping, healthcare professionals (HCPs) can offer valuable insights grounded in clinical experience and long-term therapeutic relationships. Existing studies have largely focused on parents’ perspectives, with limited attention given to those of HCPs. Exploring caregiver needs from HCPs’ perspective can therefore complement parental accounts and provide a more comprehensive understanding.

This study aims to explore the challenges and needs of parents and caregivers from the perspective of HCPs who work closely with them. Findings are discussed in relation to existing literature, integrating HCPs’ insights with caregivers’ perspectives to identify potential gaps and discrepancies in perceived needs. This study contributes to the limited body of research on ASD caregiving and offers recommendations for needs assessment and intervention development.

## Methods

This study involved in-depth interviews with HCPs providing tertiary specialist clinical services for ASD in a university-affiliated tertiary hospital in Hong Kong. Ethical approval was granted by The Joint Clinical Research Ethics Committee established by The Chinese University of Hong Kong and New Territories East Cluster (Joint CUHK-NTEC CREC, Ref: 2021.591). This report follows the Consolidated Criteria of Reporting Qualitative Studies (COREQ) guidelines [17] (Supplemental File 1).

### Participants

We purposively sampled HCPs providing tertiary specialist clinical services for children with ASD to ensure representation across roles, including psychiatrists, psychiatric nurses, clinical psychologists, occupational therapists, hospital-based teachers, and social workers. Participants were identified and recruited via a university-affiliated tertiary hospital in Hong Kong. All participants had >10 years’ experience assessing and treating children with ASD and supporting their families. Sample size was guided by the principle of information power [18].

### Data collection

Interviews were facilitated by four team members: two psychiatrists (SC and OW), a doctoral student (KM), and a research coordinator with a psychology background (OC). A topic guide developed with input from psychiatrists based on clinical experience was used to steer semi-structured discussions on diagnosis, symptom management, family dynamics, and stigma (Supplemental File 2). The guide included open-ended questions and supplementary prompts, while allowing flexibility to pursue spontaneous, in-depth discussions [19]. We conducted group interviews with psychiatric nurses who work as a team, for scheduling practicality and to enable shared reflection. Interviews were conducted in person or via videoconference and lasted 90-120 minutes. Audio recordings were transcribed verbatim; transcripts were checked for accuracy, de-identified, and stored on password-protected university drives.

### Data analyses

Interview transcripts were imported into NVivo 14 software and analysed using reflexive thematic analysis [20,21]. After familiarisation, coding proceeded inductively. Team members with backgrounds in psychology (KM) and psychiatry (SC and OW) met regularly to develop and refine codes. The lead author (KM) then collated codes and generated initial themes, which were further developed through iterative reflection and discussions with an experienced researcher in the field of child and adolescent mental health (AMB). Analysis was conducted at the latent level, attending to underlying assumptions and conceptualisations shaping participants’ accounts.

### Patient and public involvement

Stakeholders were involved in the design, conduct, and dissemination of this research. We received input from a panel of psychiatrists for the development of the interview guide. Results were disseminated in a mindfulness-based psychoeducation app coproduced with HCPs for parents of children with autism to complement existing clinical services and bridge service gaps for those who are currently on the waiting list to be seen by a psychiatrist for diagnostic assessment and clinical management.

## Results

We interviewed 16 HCPs (14 females and 2 males), including seven one-on-one interviews with two psychiatrists, two clinical psychologists, one occupational therapist, one social worker, and one hospital-based teacher, and three triad interviews with nine psychiatric nurses. Overall, three main themes were generated from our analyses: (1) Emotional strain and its negative impact on parenting practices, (2) Misunderstanding about ASD and its management, and (3) Acknowledgement and acceptance of the ASD diagnosis. Table 1 summarises the main themes and key recommendations from HCPs. These themes will be described below with illustrative quotes.

**Table 1.** Overview of themes identified and corresponding recommendations.

| Theme | Key ideas | Key recommendations |
| --- | --- | --- |
| Emotional strain and its negative impact on parenting practices | • Challenging emotions experienced around child's prognosis<br>• Emotion-driven parenting fuelling conflicts within families<br>• Harsh and unproductive parenting that may be emotionally harmful<br>• Lack of parental self-care | • Adopt low-energy approach for responses during conflicts<br>• Actively ignore child's unwanted behaviours without ignoring the child<br>• Reconnect through open discussion about emotions and concerns<br>• Prioritise self-care through mindfulness or hobbies |
| Misunderstanding about ASD and its management | • Stereotyped understanding of ASD characteristics shaped by misinformation<br>• Misinterpretation of child's behaviours and needs<br>• Misconstrued patterns of behavioural reinforcement<br>• Inconsistent boundaries and limits confusing children and straining families | • Be cautious about misinformation or misinterpretation of online information<br>• Adopt a curious and investigate mindset<br>• Conduct functional analysis of child's behaviours<br>• Use child's restricted interests as motivators for reinforcements<br>• Ensure both parents contribute equally expressing affection and setting boundaries |
| Acknowledgement and acceptance of the ASD diagnosis | • Unrealistic expectations about prognosis<br>• Excessive focus on outcomes over progress and growth<br>• Overemphasis on deficits over strengths and uniqueness<br>• Challenges to developing double-empathy across the neurodiversity spectrum<br>• Limited recognition of acceptance as a process of personal growth | • Adopting a stepwise approach<br>• Strengths-based parenting<br>• Perspective taking<br>• Third-wave cognitive behavioural therapies to support parents |

### Theme 1: Emotional strain and its negative impact on parenting practices

HCPs reported that parents often experience a range of challenging emotions in response to their child’s prognosis, including helplessness and loneliness stemming from the lack of support, hurt and guilt from the lack of validation of parenting efforts, sadness and anxiety about their child’s progress and development, as well as shame and embarrassment linked to public self-stigma around ASD. These intense and heightened emotions can influence parenting behaviours and sometimes lead to conflict within families, particularly when parents are unaware of their own emotional states.

> *Parents often want a crystal-ball approach – they want to know what their child may be able to do in the future, which causes them a lot of anxiety. (Psychiatric nurse)*

> *Parents might not have much of the warm and nurturing interactions that make parenting rewarding, hence they are often disheartened… some parents might adopt a hostile, rejecting, and detached attitude to cope. (Psychiatrist)*

Such emotional intensity may be uncomfortable and overwhelming for children sensorily and cognitively, hindering their ability to process information and potentially disrupt parent-child dynamics and relationships. HCPs suggested that supporting parents to recognise and regulate their own emotions could improve interactions, recommending low-energy responses during conflict, ignoring unwanted behaviour without ignoring the child, and reconnecting through open discussion about emotions and concerns.

> *A crucial thing parents often don’t realise is that children find it challenging when parents are in a state of emotional arousal. Children feel uncomfortable and are highly sensitive due to intensified sensory experiences. The discomfort affects their ability to receive and understand information, particularly in terms of mentalisation and communication. Children may also adopt antagonistic and paranoic attitudes, pushing others away or avoiding interactions. (Psychiatrist)*

> *It’s not productive to get angrier than the child, so we [parents] all need to pay attention to our own emotions and attune ourselves to the child… approaching the situation with an observer’s mindset and a low-energy approach can help. When the child is throwing a tantrum, they are like a tub of hot water, and if the parent is also equally emotionally charged, there will no chance for cooling down. (Psychiatrist)*

Furthermore, HCPs also noted that some parents, in moments of distress, may resort to harsh parenting that are often unproductive. For example, parents may use verbal punishment, such as shaming, mocking, or threatening abandonment when children misbehave, which are often rooted in their own childhood experiences.

> *Parents who opt for threatened abandonment often resort to hurtful and abusive language, saying things like ‘I don’t want you anymore’. This is never appropriate or effective in parenting and can have long-lasting negative effects on the child’s self-esteem, emotional wellbeing, and trust in their parents… Children often immediately show compliance when they realise that they are being ignored and revert to their usual behaviours, which is when parents should try to reconnect with them because the problematic behaviour has already been corrected. (Psychiatrist)*

> *Perhaps this sort of parenting style comes from how we were brought up, but it is so important for us [parents] to break the cycle of harmful parenting passed down through previous generations. Parents should refrain from using insults, as those memories can stay with them. (Social worker)*

These approaches were described as ineffective and potentially harmful to children’s emotional development. Finally, they emphasised the importance of parental self-care, such as mindfulness or hobbies, to maintain emotional resilience through enhancing personal capacity to reflect and destress, ultimately enabling parents to better support their children.

> *Often, it’s not that parents don’t know how to teach their children, but rather they lack the emotional energy to do so when in a state of despair. Whilst experiencing intense emotions that parents may feel like they cannot handle, practising self-care can help them turn towards acceptance… mindfulness practices can help parents find their personal space and help take care of themselves. (Psychiatrist)*

> *[Parents] taking good care of themselves also helps to improve outcomes for children. Rather than focusing on how to manage their children, the importance of having personal space to recharge and pamper oneself is often overlooked. (Clinical psychologist)*

### Theme 2: Misunderstanding about ASD and its management

HCPs reported that parents often seek information about ASD online, which can lead to misunderstandings about ASD, often shaped by misinformation or misinterpretation. HCPs noted that parents often focus on diagnostic traits and overlook individual differences in the presentation of these traits between children, as well as co-occurring conditions such as OCD and ADHD. For instance, while parents may recognise social communication difficulties, they may assume that all children with ASD are quiet or socially withdrawn, overlooking the challenges that children have in interpreting social cues and engaging in social reciprocity, especially in children who appear sociable or talkative.

> *Some people fail to understand why their child, who is overly friendly and lacks boundaries, is thought to have ASD. This is something that may not be found in the information parents search for [on the internet], but it is important for them to know. (Psychiatric nurse)*

> *It is a very common misconception as parents often think that individuals with ASD tend to and prefer to be more isolated, and we often hear them say ‘I don’t understand why the doctor said my child has ASD when he talks non-stop, annoys people all day long, and actively seeks out playmates’, but parents don’t see how the child cannot read social cues, and when people ask him to join them, he doesn’t respond or follow them. (Psychiatric nurse)*

HCPs also suggested that parents may overlook evolving traits within their own child, as familiarity with certain behaviours can shape perceptions over time. For example, repetitive behaviours are often viewed purely as symptoms of autism, rather than as strategies for managing anxiety or expressing emotion. HCPs encouraged parents to adopt a curious and investigative mindset, recognising the intentions, motivations, and mentalisation underlying their child’s behaviour.

> *Very often, when [the child] are stubborn (and attached to certain objects), it’s because that object gives them a sense of security (that cannot often be explained). They might feel that it provides a sense of safety… There is always a reason behind their stubbornness. Once the parents figure out the true reasons, they can understand how to manage the stubbornness. (Hospital-based teacher)*

> *Parents often think that their children going against them deliberately. Although this might be true at times, often children might just lack the ability to complete tasks… Parents need to learn to adopt an ‘investigative’ process of mentalisation to understand their children and their behaviours. Parenting can go a lot more smoothly if parents can carefully explore their child’s inner world and show them that they understand their thoughts and feelings. (Psychiatrist)*

Some misunderstandings were also linked to misconstrued patterns of behavioural reinforcement. HCPs had observed that parents often give more attention to negative behaviours than positive ones, unintentionally reinforcing the former. They recommended conducting functional analysis of behaviour to identify appropriate responses and using children’s restricted interests as motivators for positive reinforcement.

> *Parents need to carry out a functional analysis of the situation, carefully observe the antecedents and consequences surrounding a behaviour, whether the antecedents gradually lead to negative behaviours and whether the consequences contribute to future repetitions of such behaviours… Parents and teachers often unintentionally reinforce negative, unwanted behaviours such as hitting, pulling hair, and spitting, by reacting with great attention and strong, negative reactions that children desire to achieve. Instead, giving attention and strong, positive, and affirmative reactions when they are not engaging in these disruptive behaviours will help them extinguish such patterns of disruptive behaviours through positive reinforcement. (Clinical psychologist)*

> *Parents should not invalidate [children’s] stereotyped interests. Instead, if used appropriately, stereotyped interests can become a form of reinforcement. However, parents often tend to disregard their seemingly boring interests, leading to the emergence of negative behaviours. (Clinical psychologist)*

Conversely, overindulging repetitive interests and overaccommodating ASD-associated behaviours was described as potentially maladaptive, as it may limit the child’s skill development and disrupt family dynamics. These patterns can also negatively impact neurotypical siblings, who may receive less attention or be expected to unfairly adapt to the needs of their sibling with ASD.

*Very often, the typically developing siblings are very understanding and comforting. But because of this, parents always say things like ‘Grow up, understand your brother [with ASD] and accommodate his needs.’ Whilst every mother claims that they are not biased, isn’t this not a form of favouritism? (Psychiatrist)*

HCPs emphasised the need to set boundaries and limits that is consistently maintained across caregivers. Inconsistent approaches were said to confuse children, as they may struggle to understand, learn, and integrate rules, and strain family relationships, as children may gravitate towards the more lenient parent. HCPs recommended that parents align their caregiving approaches and share responsibilities, ensuring that both parents contribute equally to expressing affection and setting boundaries.

> *Very often what we observe is that fathers take up the role of playing and rewarding, while mothers manage everything, such as children’s studies, daily routines, and reminders. (Psychiatrist)*

> *This division of labour can be confusing for children and make things more challenging, as children may lean towards the parent who is easier for them behaviourally, and it will be difficult for them to learn any good habits. (Psychiatrist)*

### Theme 3: Acknowledgement and acceptance of the ASD diagnosis

HCPs observed that some parents struggle to come to terms with their child’s ASD diagnosis, which may manifest as denial, reluctance to seek help, or unrealistic expectations about prognosis based on stereotypes and norms.

> *Parents are often resistant to the idea [of their children having autism], and only reluctantly acknowledge their child’s autism after going through a series of failed attempts to address the challenges as conventional methods prove ineffective. (Psychiatric nurse)*

> *Sometimes parents do come to us and ask us ‘When will they get better? When will they become normal again?’. In roughly half of the cases I see [in the clinic], parents are still looking at neurotypical children, which brings them more sadness. (Psychiatric nurse)*

> *Don’t expect them [children with autism] to be extremely flexible, just aim for a certain level of adaptability, with some unique qualities, and that will be enough. At the very least, aim for the child to be able to live a functional life, and will not at a stage where they are unable to function at all. (Psychiatrist)*

This often leads to an excessive focus on outcomes, viewing deficits as problems to be fixed rather than recognising children’s capabilities and progress. Such outcome-focused approaches may undermine a child’s motivation, provoke strong emotional reactions, and strain parent-child relationships.

> *Parents tend to focus on ‘fixing’ children’s problems/issues. However, it is important for them to reflect on more positive aspects and appreciate the progress of improvements. (Psychiatric nurse)*

> *Children can feel it when their parents cannot see their good side and might begin to perceive themselves as inadequate. As a result, they may be reluctant to seek help, fearing that others will discover their flaws and overlook their strengths. (Psychiatric nurse)*

Instead, HCPs encouraged a stepwise, incremental approach, focusing on small improvements, which can foster a sense of success and mastery in the child, promote positive emotions, and support longer-term adaptive functioning.

> *Parents should appreciate progress and adopt an incremental approach – for example, ‘ask them to try to write three lines, and if they expressed difficulties, ask for two instead and accept that for the day’. (Hospital-based teacher)*

> *It might be helpful for parents to try to see their child’s progress and improvements by looking back to the child’s state a year ago and comparing with the current state… this way it is easier for parents to recognise and appreciate the progress and be hopeful for the improvements that are yet to come. (Psychiatric nurse)*

> *If parents can appreciate these patterns in their children, it might help parents have a slightly greater sense of satisfaction or a higher level of acceptance. (Psychiatrist)*

Acceptance was described as essential for shifting towards strengths-based parenting, enabling children with ASD to feel acknowledged and valued, which can be beneficial for children’s self-perception and self-confidence, and enhancing parents’ own sense of satisfaction. HCPs encouraged parent to acknowledge and praise their child’s efforts and qualities, which is particularly important in cultures where emotions are conveyed less directly, and societies where the neurodiversity approach is not yet widely recognised or established.

> *Especially in our culture, families may tend to express emotions indirectly, making it challenging to convey thoughts and feelings with words… however, compliments can go a long way, and can help children feel acknowledged and valued. (Psychiatric nurse)*

> *It’s not just them [children with ASD] not being able to empathise – we may also not be able to empathise with them too, as we may never understand why they are so averse to certain noises. However, it is important to be respectful and try considering things from their perspectives too. (Psychiatrist)*

HCPs also highlighted the importance of perspective-taking in deepening parents’ understanding of their child’s intentions and behaviours, echoing the ‘double empathy problem’ often discussed in the neurodiversity movement, which emphasises the need to bridge misunderstandings between ASD and non-ASD individuals. Although acceptance can be challenging, HCPs highlighted that it is achievable with appropriate guidance and resources to support parents, such as third-wave cognitive behavioural therapies for parents.

> *It might not be easy to find positive aspects in negative situations, but seeing the situation from the child’s perspective can help parents find opportunities to praise behaviours that may be overlooked or misinterpreted. (Psychiatric nurse)*

> *Even when parents say that they have accepted it [their children’s ASD diagnoses], it doesn’t always mean true acceptance. Acceptance is a process, which includes awareness, acknowledgement, and accommodation. Accept, then find committed actions to change. Therapies, such as cognitive behavioural therapy or acceptance and commitment therapy, can be helpful. (Clinical psychologist)*

## Discussion

To our knowledge, this is the first study to explore the challenges and needs of parents and caregivers of children with ASD from the perspectives of HCPs. Drawing on their expertise and long-term therapeutic relationship with families, HCPs offered a holistic view of parenting challenges that caregivers themselves may not always recognise. This perspective extends the predominantly caregiver-focused literature, providing valuable insights and practical recommendations for supporting families and addressing a significant gap in the evidence base on caregiving in the context of ASD.

In this study, HCPs highlighted how parents’ and caregivers’ emotional distress can have downstream effects on parenting practices, and in turn, negatively affect children’s development. This complements previous studies exploring how ASD symptoms and challenging behaviours contribute to parental distress [22] and aligns with recent evidence linking parental stress and depression to parenting efficacy [23], parenting stress to parenting practices [24], and caregiver strain to externalising problem behaviours in children with ASD [25]. HCPs also emphasised the importance of acknowledging and accepting the diagnosis, echoing previous research showing that internalised stigma surrounding ASD can harm their parental emotional wellbeing [26,27]. To facilitate acknowledgement and acceptance, professionals advocated for strengths-based parenting, which benefits both children’s development and parents’ satisfaction, consistent with growing interest in such approaches within interventions and education [28,29]. The recommendation to use third-wave cognitive behavioural therapies to support parents on their journey of acceptance, aligns with findings that mindfulness and self-compassion can buffer the negative impact of stigma [30,31], and reduce parental stress [32,33].

In addition to emotional strain, HCPs drew attention to how misinformation can lead to misunderstanding about ASD, adversely influencing parenting practices. This insight may help explain inconsistent findings in the literature, as a recent systematic review found no indication that misunderstanding was an issue in the pre-diagnostic phase [34], whereas a scoping review reported a lack of knowledge about ASD as a key concern during diagnosis [7]. This discrepancy may reflect the broad conceptualisation of “understanding”, in which misunderstanding is often conflated with lack of knowledge [35]. The distinction is important: while both may appear as insufficient knowledge, especially given the widespread use of unidimensional measures of ASD knowledge [36], misunderstanding involves false beliefs perceived as accurate, which can shape parenting responses and behaviours in different ways. HCPs’ accounts of misconstrued behavioural reinforcement are consistent with previous reports of “reinforcement traps” in family processes in the general population [37], but appear exacerbated in ASD due to the prevalence of rituals and repetitive behaviours. This represents a novel contribution, as these dynamics are less likely to emerge from caregiver only perspectives.

Across all themes, HCPs provided a range of recommendations to support parents and caregivers of children with ASD in addressing these challenges. Central to these recommendations is the overarching emphasis on relational skills. They encouraged parents to take their children’s perspective and adopt a curious and investigative mindset, exploring the intentions, motivations, and mentalisation underlying their child’s behaviour. HCPs also emphasised open communication, consistent attention, and the importance of recognising siblings’ experiences, an area increasingly addressed in the literature [38,39]. These recommendations could inform psychoeducation in routine clinical practice and the development of family-centred interventions to reduce the bidirectional cycle of parental stress and child behavioural and emotional difficulties. Future research should investigate whether enhancing parents’ relational skills improves parent-child interactions, parental wellbeing and child behavioural outcomes.

Finally, although the prevalence of ASD in Hong Kong is comparable to that in Western countries [40], findings may not reflect the perspectives of health professionals in other countries, particularly where healthcare provision for autism differs considerably. While the qualitative design enabled an in-depth exploration of experience and produced rich and detailed data, we were unable to explore causal relationships or mechanisms, and future mixed-methods research should examine relationships between caregiver skills and wellbeing.

## Conclusions

This study identified caregiving challenges and needs that have previously been less studied, including misunderstandings about ASD and strategies for managing children’s behaviours and symptoms. HCPs emphasised the importance of building relationships with children with ASD through curiosity, perspective-taking, appreciation of strengths, and expressions of affection. Incorporating HCPs’ perspectives revealed aspects of caregiving that parents and caregivers may not always recognise, thereby broadening the evidence-base on caregiving in ASD. These findings highlight the importance of family-centred care and point to potential loci for psychoeducation to better support parents and caregivers of ASD in addressing caregiving challenges.

## Statement and Declarations

## Acknowledgements

The authors would like to thank our professional acquaintances at the university-affiliated tertiary hospital in Hong Kong for their referral and participation in this study. We would also like to thank medical students and copywriter engaged by The Chinese University of Hong Kong for their assistance in interview transcription.

## Author contributions

All authors contributed to the study conception and design, material preparation, and data collection. KM, OC, OW, and SC collected the data. KM conducted the analysis with contributions from AMB. KM wrote the first draft of the manuscript. All authors read and approved the final manuscript.

## Patient consent for publication

Not applicable

## Ethical approval

This study was approved by The Joint Clinical Research Ethics Committee established by The Chinese University of Hong Kong and New Territories East Cluster (Joint CUHK-NTEC CREC, Ref: 2021.591). Informed consent was obtained from all participants before taking part in the study.

## Data availability statement

Data are available upon reasonable request.

## Supplemental material

Supplemental material for this article is available online.

## References

1 Steinhausen H-C, Mohr Jensen C, Lauritsen MB. A systematic review and meta-analysis of the long-term overall outcome of autism spectrum disorders in adolescence and adulthood. Acta Psychiatr Scand. 2016;133:445–52. doi: 10.1111/acps.12559

2 Bonis S. Stress and Parents of Children with Autism: A Review of Literature. Issues Ment Health Nurs. 2016;37:153–63. doi: 10.3109/01612840.2015.1116030

3 Van Steijn DJ, Oerlemans AM, Van Aken MAG, et al. The reciprocal relationship of ASD, ADHD, depressive symptoms and stress in parents of children with ASD and/or ADHD. J Autism Dev Disord. 2014;44:1064–76. doi: 10.1007/S10803-013-1958-9

4 Long CE, Gurka MJ, Blackman JA. Family Stress and Children’s Language and Behavior Problems: Results From the National Survey of Children’s Health. Topics Early Child Spec Educ. 2008;28:148–57. doi: 10.1177/0271121408318678

5 Depape AM, Lindsay S. Parents’ experiences of caring for a child with autism spectrum disorder. Qual Health Res. 2015;25:569–83. doi: 10.1177/1049732314552455

6 Wong V, Yu Y, Keyes ML, et al. Pre-diagnostic and Diagnostic Stages of Autism Spectrum Disorder: A Parent Perspective. Child Care in Practice. 2017;23:195–217. doi: 10.1080/13575279.2016.1199537

7 Makino A, Hartman L, King G, et al. Parent Experiences of Autism Spectrum Disorder Diagnosis: a Scoping Review. Review Journal of Autism and Developmental Disorders 2021 8:3. 2021;8:267–84. doi: 10.1007/S40489-021-00237-Y

8 Marsh A, Spagnol V, Grove R, et al. Transition to school for children with autism spectrum disorder: A systematic review. World J Psychiatry. 2017;7:184. doi: 10.5498/WJP.V7.I3.184

9 Vohra R, Madhavan S, Sambamoorthi U, et al. Access to services, quality of care, and family impact for children with autism, other developmental disabilities, and other mental health conditions. Autism. 2014;18:815–26. doi: 10.1177/1362361313512902

10 O’Nions E, Happé F, Evers K, et al. How do Parents Manage Irritability, Challenging Behaviour, Non-Compliance and Anxiety in Children with Autism Spectrum Disorders? A Meta-Synthesis. J Autism Dev Disord. 2018;48:1272–86. doi: 10.1007/S10803-017-3361-4

11 Kinnear SH, Link BG, Ballan MS, et al. Understanding the Experience of Stigma for Parents of Children with Autism Spectrum Disorder and the Role Stigma Plays in Families’ Lives. J Autism Dev Disord. 2016;46:942–53. doi: 10.1007/S10803-015-2637-9

12 Salleh NS, Abdullah KL, Yoong TL, et al. Parents’ Experiences of Affiliate Stigma when Caring for a Child with Autism Spectrum Disorder (ASD): A Meta-Synthesis of Qualitative Studies. J Pediatr Nurs. 2020;55:174–83. doi: 10.1016/J.PEDN.2020.09.002

13 Bekhet AK, Johnson NL, Zauszniewski JA. Resilience in Family Members of Persons with Autism Spectrum Disorder: A Review of the Literature. Issues Ment Health Nurs. 2012;33:650–6. doi: 10.3109/01612840.2012.671441

14 Karst JS, Vaughan A, Hecke V. Parent and Family Impact of Autism Spectrum Disorders: A Review and Proposed Model for Intervention Evaluation. Clin Child Fam Psychol Rev. 2012;247–77. doi: 10.1007/s10567-012-0119-6

15 de Leeuw A, Happé F, Hoekstra RA. A Conceptual Framework for Understanding the Cultural and Contextual Factors on Autism Across the Globe. Autism Research. 2020;13:1029–50. doi: 10.1002/AUR.2276

16 Ademosu T, Ebuenyi I, Hoekstra RA, et al. Burden, impact, and needs of caregivers of children living with mental health or neurodevelopmental conditions in low-income and middle-income countries: a scoping review. Lancet Psychiatry. 2021;8:919–28. doi: 10.1016/S2215-0366(21)00207-8

17 Tong A, Sainsbury P, Craig J. Consolidated criteria for reporting qualitative research (COREQ): a 32-item checklist for interviews and focus groups. International Journal for Quality in Health Care. 2007;19:349–57. doi: 10.1093/INTQHC/MZM042

18 Malterud K, Siersma VD, Guassora AD. Sample Size in Qualitative Interview Studies: Guided by Information Power. Qual Health Res. 2016;26:1753–60. doi: 10.1177/1049732315617444

19 Ryan F, Coughlan M, Cronin P. Interviewing in qualitative research: The one-to-one interview. Int J Ther Rehabil. 2009;16:309–14. doi: 10.12968/IJTR.2009.16.6.42433

20 Braun V, Clarke V. Using thematic analysis in psychology. Qual Res Psychol. 2006;3:77–101. doi: 10.1191/1478088706QP063OA

21 Braun V, Clarke V. Reflecting on reflexive thematic analysis. Qual Res Sport Exerc Health. 2019;11:589–97. doi: 10.1080/2159676X.2019.1628806

22 Siu QKY, Yi H, Chan RCH, et al. The Role of Child Problem Behaviors in Autism Spectrum Symptoms and Parenting Stress: A Primary School-Based Study. J Autism Dev Disord. 2019;49:857–70. doi: 10.1007/S10803-018-3791-7

23 Giallo R, Wood CE, Jellett R, et al. Fatigue, wellbeing and parental self-efficacy in mothers of children with an Autism Spectrum Disorder. Autism. 2013;17:465–80. doi: 10.1177/1362361311416830

24 Suvarna V, Farrell L, Adams D, et al. Differing relationships between parenting stress, parenting practices and externalising behaviours in autistic children. Autism. 2025;29:711–25. doi: 10.1177/13623613241287569

25 Lindly OJ, Shui AM, Stotts NM, et al. Caregiver strain among North American parents of children from the Autism Treatment Network Registry Call-Back Study. Autism. 2022;26:1460–76. doi: 10.1177/13623613211052108

26 Chan KKS, Lam CB. Self-stigma among parents of children with autism spectrum disorder. Res Autism Spectr Disord. 2018;48:44–52. doi: 10.1016/J.RASD.2018.01.001

27 Mak WWS, Kwok YTY. Internalization of stigma for parents of children with autism spectrum disorder in Hong Kong. Soc Sci Med. 2010;70:2045–51. doi: 10.1016/J.SOCSCIMED.2010.02.023

28 Lee EAL, Black MH, Falkmer M, et al. “We Can See a Bright Future”: Parents’ Perceptions of the Outcomes of Participating in a Strengths-Based Program for Adolescents with Autism Spectrum Disorder. J Autism Dev Disord. 2020;50:3179–94. doi: 10.1007/S10803-020-04411-9

29 Taylor EC, Livingston LA, Clutterbuck RA, et al. Psychological strengths and well-being: Strengths use predicts quality of life, well-being and mental health in autism. Autism. 2023;27:1826–39. doi: 10.1177/13623613221146440

30 Wong CCY, Mak WWS, Liao KYH. Self-Compassion: a Potential Buffer Against Affiliate Stigma Experienced by Parents of Children with Autism Spectrum Disorders. Mindfulness (N Y). 2016;7:1385–95. doi: 10.1007/S12671-016-0580-2

31 Yip CCH, Chan KKS. Longitudinal impact of public stigma and courtesy stigma on parents of children with autism spectrum disorder: The moderating role of trait mindfulness. Res Dev Disabil. 2022;127. doi: 10.1016/J.RIDD.2022.104243

32 Li SN, Chien WT, Lam SKK, et al. Effectiveness of parent-focused interventions for improving the mental health of parents and their children with autism spectrum disorder: A systematic review and meta-analysis. Res Autism Spectr Disord. 2024;114:102389. doi: 10.1016/J.RASD.2024.102389

33 Juvin J, Sadeg S, Julien-Sweerts S, et al. A Systematic Review: Acceptance and Commitment Therapy for the Parents of Children and Adolescents with Autism Spectrum Disorder. J Autism Dev Disord. 2022;52:124–41. doi: 10.1007/S10803-021-04923-Y

34 Solia D, Albarqouni L, Stehlik P, et al. Parent concerns prior to an assessment of autism spectrum disorder: A systematic review. Autism. 2025;29:838–49. doi: 10.1177/13623613241287573

35 Shilubane H, Mazibuko N. Understanding autism spectrum disorder and coping mechanism by parents: An explorative study. Int J Nurs Sci. 2020;7:413–8. doi: 10.1016/J.IJNSS.2020.08.003

36 Harrison AJ, Slane MM, Hoang L, et al. An international review of autism knowledge assessment measures. Autism. 2017;21:262–75. doi: 10.1177/1362361316638786

37 Scott S, Dadds MR. Practitioner Review: When parent training doesn’t work: theory-driven clinical strategies. Journal of Child Psychology and Psychiatry. 2009;50:1441–50. doi: 10.1111/J.1469-7610.2009.02161.X

38 Watson L, Hanna P, Jones CJ. A systematic review of the experience of being a sibling of a child with an autism spectrum disorder. Clin Child Psychol Psychiatry. 2021;26:734–49. doi: 10.1177/13591045211007921

39 Burnham Riosa P, Ensor R, Jichici B, et al. How my life is unique: Sibling perspectives of autism. Autism. 2023;27:1575–87. doi: 10.1177/13623613221142385

40 Wong OW, Chan SS, Chau SW, et al. Autism epidemiology in Hong Kong children and youths aged 6–17: Implications on autism screening and sex differences in the community. Autism. Published Online First: 27 July 2025. doi: 10.1177/13623613251360269

