## Supplementary material for "Healthcare professionals’ perspectives on the challenges faced by parents of children with autism and recommendations to address them: A qualitative study": Topic Guide (from another version of this preprint: version 1, 2025-11-15)

**Supplemental Material**

Topic guide for semi-structured interviews.

| Topic | Example of questions and probes |
| --- | --- |
| Diagnosis | - What was it like for parents when they first got informed about their child’s ASD diagnosis? How do they react and/or feel? - What are parents’ own perception of their child’s diagnosis? |
| Symptom management | - What are parents’ main difficulties in caregiving with regards to their child’s ASD symptoms? Why is that the case? - How do they cope/respond to these situations? - What do parents need the most in order to overcome these difficulties? |
| Family dynamics | - How does the wider family dynamics affect parents’ experience of caregiving for children with ASD? - Are there any factors or situations that are particularly helpful or causing extra stress? |
| Stigma | - What are parents’ experiences of stigma related to ASD? - What are their thoughts and feelings when faced with public stigma? |
